# Factors associated with commuting stress among tertiary education employees in Georgetown, Guyana

**DOI:** 10.1101/2024.02.23.24303247

**Authors:** Davon Van-Veen, Hue-Tam Jamme, Heather Ross, Netra Chhetri

## Abstract

The aim of this study was to identify factors associated with commuting stress using symptomatology among tertiary education employees in Georgetown Guyana. A cross sectional survey was conducted among 427 (100 male, 317 female) participants, with a mean age of 29.6(sd=8.7) years. Data about their demographic characteristics, psychosocial measures (perceived stress, coping and resilience), characteristics about their commute (length, mode, and satisfaction with commute infrastructure), and how often they experienced selected symptoms associated with stress on the daily commute was collected using a self administered questionnaire. A commuting stress score was calculated for each participant and this was followed by regression analysis. The results showed that after controlling for resiliency, coping, perception of job, total life stress, income levels and education all of which can contribute to how persons perceive hassles in their lives, the regression model demonstrated that females, persons who used public transportation, persons who have longer commute times and persons who are less satisfied with the commute infrastructure are more likely to report that they experience symptoms associated with stress while engaged in the daily commute compared to males, persons who use private transportation, persons who have shorter commute times and persons who are more satisfied with the commute infrastructure respectively. Once all controlled variables were accounted for, it appears that for this study population, age, where they live and whether or not they actively or passively commuted did not have any meaningful impact on whether they would report if they experienced stress associated symptoms while engaged in the daily commute. he results suggest the need for a multi-pronged approach to address commuting stress, involving individual healthcare, mental-health-centric urban transport design, improved commute infrastructure, and increased employer engagement. Future studies employing qualitative and experimental methods are recommended.

## Introduction

The majority of contemporary urbanisation, a complex social and economic process that occurs through various forms of urban growth and expansion drastically altering the social and physical environment in which people live and operate, is occurring in low and middle income countries(LMICS).[1,2] Not surprisingly these changes can have a significant impact on both physical and mental health on urban inhabitants.[3–5] Increased access to modern technology in the urban environment has induced a fast-paced life which is also characterised by a heavy workload, fast paced lifestyle, communication, quick decisions, multitasking, pressure to be active on social media, and high mobility, all of which have a direct impact on people’s health [6,7] however for the purposes of this paper, there is going to be an exclusive focus on high mobility.

The majority of mobility in the urban environment consists of the daily commute which appears to be intensifying in both the developed and developing world. This intensification is a result of an increase in both the frequency and length/duration of commutes to work.[8,9] There is a significant body of literature that examines how the increased intensity of commuting *to* and *from* work can have detrimental effects on health. Impacts of lengthy commutes include an increased incidence of respiratory illnesses, muscular injuries, and reduced physical activity along with the associated undesirable health outcomes.[10–14] Additionally, an emerging body of scholarship reveals that a lengthy commute to workplaces is also associated with poor mental health which may be expressed as stress.[15–19] The commute is more often than not a daily activity, thus if it is stressful, a person may be placed under stress on an ongoing basis.

Stress is the physiological and psychological response to a situation that a person has perceived to be negative and has made a determination that they do not have the resources to address the problem, accounting for all other factors.[20–22] In addition to being an indicator of poor mental health, chronic exposure to stress is also associated with many chronic non-communicable diseases (CNCDs) including but not limited to diabetes, cardiovascular and cerebrovascular pathologies, chronic obstructive pulmonary disease, and malignant neoplasms.[23] CNCDs continue to be a significant public health concern globally, accounting for most of the reported global mortality and morbidity with the global burden rapidly shifting towards the LMICs.[24–26] Furthermore, long-term exposure to stress is more likely to be in a state of constant anger and irritability, leading to strained personal relationships which can further lead to to potential isolation, anxiety, and depression. If it is severe enough, it may even lead to suicide.[27–29]

The question therefore becomes who is more likely to experience commuter stress and there have been a few studies that attempted to examine these factors. For example, women are more likely to experience commuter stress than men.[30–34] Similarly, active commuters are less likely to experience stress than passive commuters.[35–37] The issue however with the majority of these studies either examined only the intrinsic factors (factors related to the commuter) or the extrinsic factors (factors related to the commute process). Also, almost all of the previous studies were conducted in economies that were considered fully urbanised (that is, more than 50% of their population live in urban agglomerations) and there has been a dearth of studies that examined this issue in economies that are in transition to urbanisation. Also, the majority of previous studies used only Likert or binary scales to examine stress experienced during the commute. They did not focus on the physical and psychological symptoms of stress. In order to address the preceding gaps identified, this paper aims to to identify what factors (both extrinsic and intrinsic) are associated with experiencing stress while engaged in the daily commute using in an emerging economy that has less than 50% of its population living in an urban environment using symptomatology to measure commuting stress.

The next section of this paper detailed the methods This is followed by the presentation of the results which consists of the presentation of the sample characteristics, the individual risk factor assessment and regression models that include all of the factors that were tested. These results and the implications with respect to the literature are then discussed in the next section after which some final conclusions along with some recommendations are made.

## Method

### Study design and setting

This observational cross-sectional study was conducted in Georgetown, the capital city of Guyana, between March 1st to April 30th, 2023. As the country’s political and economic centre, it serves as a critical hub for administrative, commercial, and cultural activities. While the reliable data on the population of Georgetown is limited, the National Census of 2012 puts its population to be around 120,000 people.

### Study Participants

Participants were drawn from among employees at the University and the University’s affiliate partners. The University of Guyana serves as the National University of Guyana and has just about 8,000 students and employs about 1,000 teaching and non-teaching staff. Universities are normally microcosms of the wider society in which they operate, therefore it is to be expected that the demographic makeup of the university would be similar to that of the society in which they function.[38–39]

Using an online sample size calculator, (Raofost, 2004), assuming an unknown population a sample size of 377 was recommended. Assuming that all questionnaires would not be returned or only partially completed, it was determined that 33% more questionnaires should be distributed therefore 505 questionnaires were distributed. To be included in the study, participants must work in Georgetown for at least 30 hours per week at an in person job. The participant must also be at least 18 years of age and have worked on the job for at least 6 months.

Convenience sampling was used until the desired sample size was achieved. Employees would be approached and given a short presentation about the rationale, objectives, and significance of the study along with guarantees that no identifiable information will be collected and that all information collected would remain confidential. After the employee gave their verbal consent to participate in the study, each employee was informed that there were three different ways to participate in the study: a shared link, a tablet with the questionnaire on it, or a physical paper questionnaire and it was for them to choose the method that they were most comfortable with. Once a choice was made, the 83-item questionnaire was made available using the medium of their choice, and the employee was thanked for their time.

### Variables

The dependent variable commuting stress was evaluated using symptomatology. This was done by asking participants to report the frequency in which they experienced a variety of symptoms associated with stress while engaged in the daily commute. These included physical symptoms such as muscle pain, headaches, nausea, difficulty breathing, fatigue, chest pain, indigestion, dizziness, sweating and constipation/diarrhoea and psychological symptoms such as feeling irritable, overwhelmed, anxious, having uncontrollable thoughts, depressed, a sense of dread, lonely, pressured, and distressed. For each symptom, they were asked whether it was experienced very often, fairly often, sometimes, almost never, or never. Based on the response a score was assigned to each symptom with very often=4 and never=1. A total commuting stress score was then generated.

The independent variables aimed to assess personal (intrinsic) factors, commute specific factors, and built environment factors. The demographic factors which were tested include age, sex, place of residence (whether they lived in the city or the suburbs), and hours of work per day. Other intrinsic factor included was the perceived quality of sleep which was measured on a five-point Likert scale, the general life stress which was measured using the Cohen’s Perceived Stress Scale (PSS-10), and how hassled a person perceived their job to be, which was measured using a sub-scale of the Daily Hassles and Mood Questionnaire (DHMQ). The DHMQ is a self-report measure developed by Dr. Thomas W. Kamarck and his colleagues in 1991, which assesses the daily hassles, mood, and health status of individuals. The job sub-scale consists of seven questions that aimed to elicit how people felt about their fellow workers, their supervisors, and their workload. It was scored from 0-21. Higher scores were associated with negative feelings about the job. The DHMQ has been found to be a valid and reliable measure, with high levels of internal consistency and test-retest reliability. It has been used in various populations, including healthy adults, medical patients, and college students. The scale has been found to be positively associated with a variety of health outcomes, such as cardiovascular disease, hypertension, and depression.[40–42] The Perceived Life Stress measured by Cohen’s Perceived Stress Scale (PSS-10) is another independent variable. The PSS-10 is a 10-item version of the scale, which was developed by Sheldon Cohen and his colleagues in 1983. The scale assesses the extent to which respondents find their lives unpredictable, uncontrollable, and overloaded in the last month. Respondents are asked to rate the degree to which they agree with each statement on a 5-point Likert scale, where 0 = never and 4 = very often. The PSS-10 has been found to be a valid and reliable measure of perceived stress, with high levels of internal consistency and test-retest reliability. In addition, the scale has been validated in various populations including, but not limited to, healthy adults, medical patients, and college students. It has been found to be positively associated with a variety of health outcomes, including cardiovascular disease, hypertension, and depression.[43–45]

The participants’ coping capabilities were the third mediating variable that was tested using the Brief Resilient Coping Scale (BRCS). This is a 4-item unidimensional scale that is designed to capture tendencies to cope with stress in a highly adaptive manner. The scale ranges from 0-20 and the higher the score, the greater the coping capabilities of the person This instrument has also been successfully subjected to multiple reliability and validity evaluations.[46–48]

Resiliency refers to the ability to adapt despite encountering less-than-ideal circumstances. This was measured by using the Brief Resilience Scale (BRS) which is a 6-point unidimensional scale that does not take into account external factors. [49] A person who is resilient tends to demonstrate high levels of emotional stability, self-efficacy, and sociability and thus better able to handle life stresses. The BRS score ranges from 1-5 with the higher values indicating higher levels of resilience. Like the BCRS, the BRS was also subject to multiple reliability and validity testing. [50–52]

The first commute-specific factors which were tested were the method of commuting whether it was active or passive, or where commuting was done by public or private means. The second commute-specific factor that was tested was the length of the commute.The study used 5-point Likert Scale questions to assess commuters’ satisfaction with various built environment factors. These factors included greenery along the route, frequency of traffic congestion, and road-use compliance by others. Additionally, satisfaction with transportation infrastructure was gauged through multiple aspects like road conditions, street lighting, and public transport. A composite score ranging from 9-45 was generated, with higher scores indicating greater satisfaction.

### Data analysis

All data were analysed using Stata Version 17.0. Descriptive analysis was performed on all variables. Non numerical data was converted to binary. For gender, males were assigned a value of 0 and females a value of 1. Those who lived in the city were assigned a value of 1 while those who lived in the suburbs were assigned a value of 0. Persons who used active and private commute modes were assigned a value of 0 while persons who used active and public forms of transportation were assigned a value of 1. Regression analysis was conducted. All values are reported with a 95% confidence interval. A *p*-value of less than 0.05 was considered to be statistically significant.

### Ethical Issues

For each participant, the purpose of the study was explained. They were informed that participation in this study was completely voluntary and that there were no consequences whether good or bad if they chose to participate in the study. They were also informed that they do not have to answer any question they feel uncomfortable about asking and they can chose to terminate their participation in the study at any point. Participants were also informed that no personally identifying data will be collected and therefore there was no way to trace a particular submission to any one participant. This information was repeated on the cover of the questionnaire that was provided. The first question of the questionnaire asked whether or not they agreed to participate in the study.

This study was approved by the Institutional Review Board of Arizona State University. (Protocol number: STUDY00017507)

## Results

Out of 505 questionnaires initially distributed, several were excluded from the final analysis for various reasons: 22 had over 10% of questions unanswered; 24 were completed by individuals working fewer than 30 hours per week; 2 were filled out by minors under the age of 18; 29 were from respondents who neither worked or lived in the study area; and 1 came from a remote worker. This left 427 valid questionnaires for final analysis There participants were relatively diverse with respect to gender, age, relationship status, ethnicity, religious affiliation, educational levels, length of commute,total life stress levels and where they resided. Table 1 shows the nominal characteristics of the participants included in the final analysis while table 2 provides a summary of the numerical characteristics.

**Table 1:** Summary of nominal characteristics of study participants (n=427)

| Parameter | Number (%) |
| --- | --- |
| **Gender** |  |
| Male | 100 (25.6) |
| Female | 317 (74.4) |
| **Relationship status** |  |
| Committed Relationship | 205 (48.2) |
| Single | 187 (44.1) |
| No response | 35 (8.2) |
| **Income level** |  |
| Low income | 280 (65.5) |
| High and middle income | 147 (34.5) |
| **Ethnicity** |  |
| Indigenous | 3 (0.7) |
| African Guyanese | 153 (36.0) |
| East Indian Guyanese | 121 (28.5) |
| Mixed descent | 148 (34.8) |
| No Response | 2 (0.5) |
| **Religious affiliation** |  |
| Christian | 311 (73.3) |
| Hindu | 64 (15.1) |
| Muslim | 29 (6.8) |
| Other/ Non-affiliated | 23 (5.4) |
| **Educational Level Completed** |  |
| Tertiary | 363 (85.2) |
| Non-Tertiary | 64 (14.8) |
| **Residence** |  |
| In Georgetown (main city) | 149 (38.8) |
| Outside of Georgetown | 278 (61.2) |
| **Commute Mode (Private/Public)** |  |
| Private | 240 (56.2) |
| Public | 187 (43.8) |
| **Commute Mode** |  |
| Active | 23 (5.4) |
| Non-Active | 404 (94.6) |

**Table 2:**
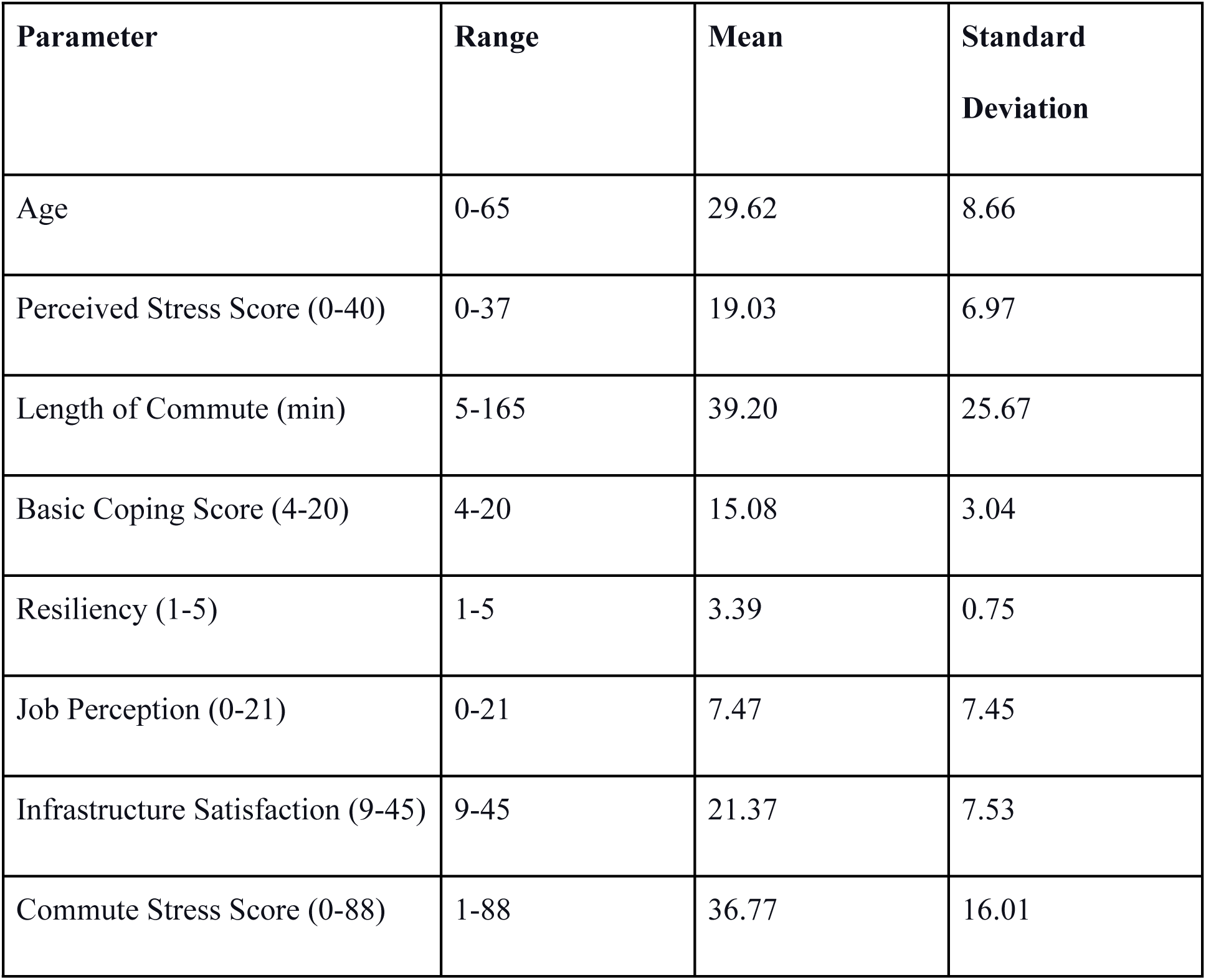
Summary of numerical characteristics of study participants (n=427)

The three most frequently reported symptoms associated with stress while engaged in the daily commute are complaints about feeling fatigued, excessive sweating and feeling hassled.

The four least commonly reported symptoms are feelings of indigestion, chest pains, dizziness and bowel dysfunction. Figure 1 shows the distribution of all the symptoms examined in this study.

**Figure 1:**
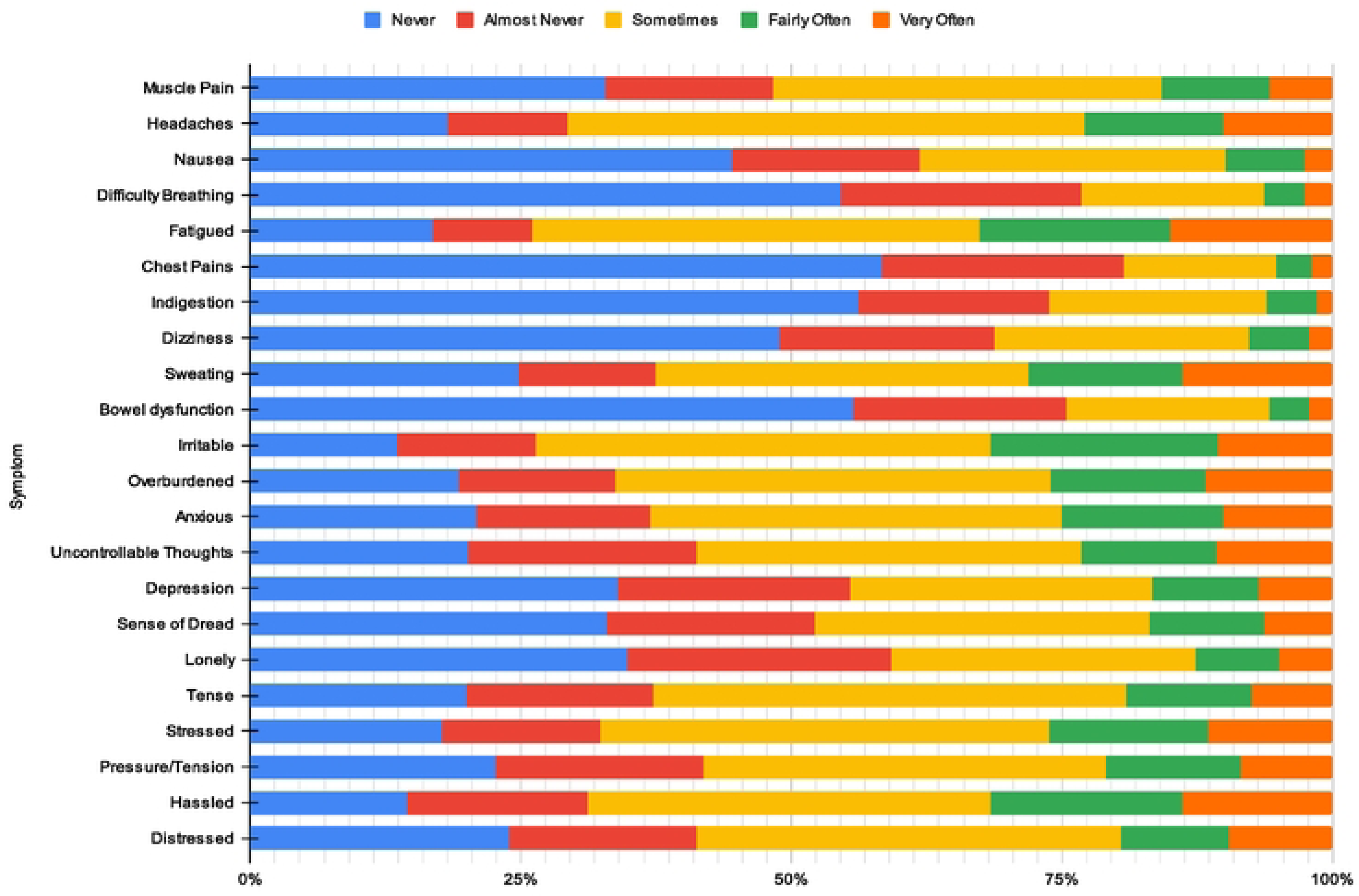
Distribution of symptoms experienced by respondents.

The results in Table 3 shows that after controlling for resilience, coping, perceived life stress, satisfaction with job, income levels and educational attainment, commuting stress levels measured using symptomatology were significantly higher in females, persons who used public means of transportation, persons with longer commute times and persons who were not satisfied with commute infrastructure. Age and location of residence appear to have no effect on the level of commuting stress as measured using symptomatology.

**Table 3:**
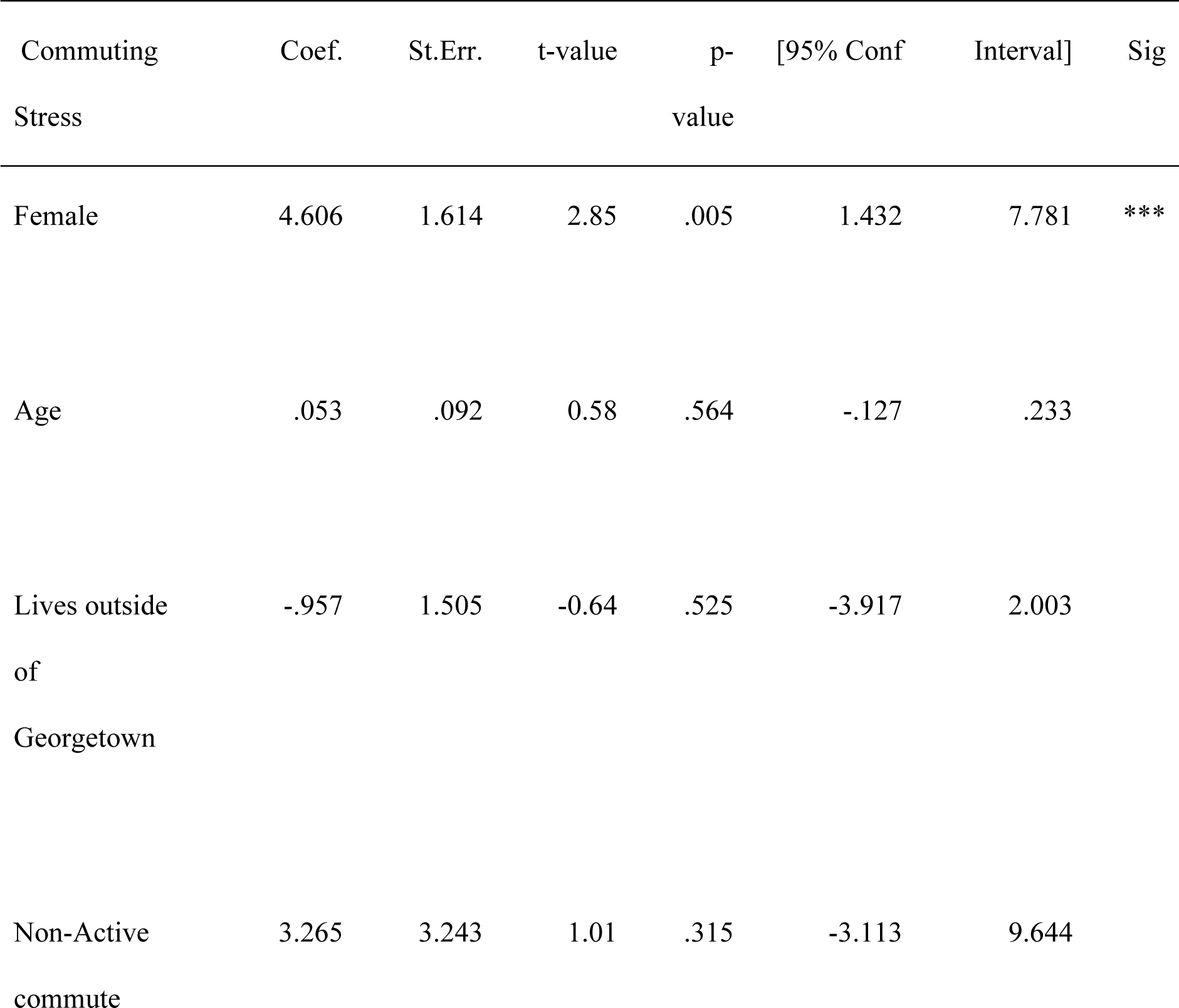
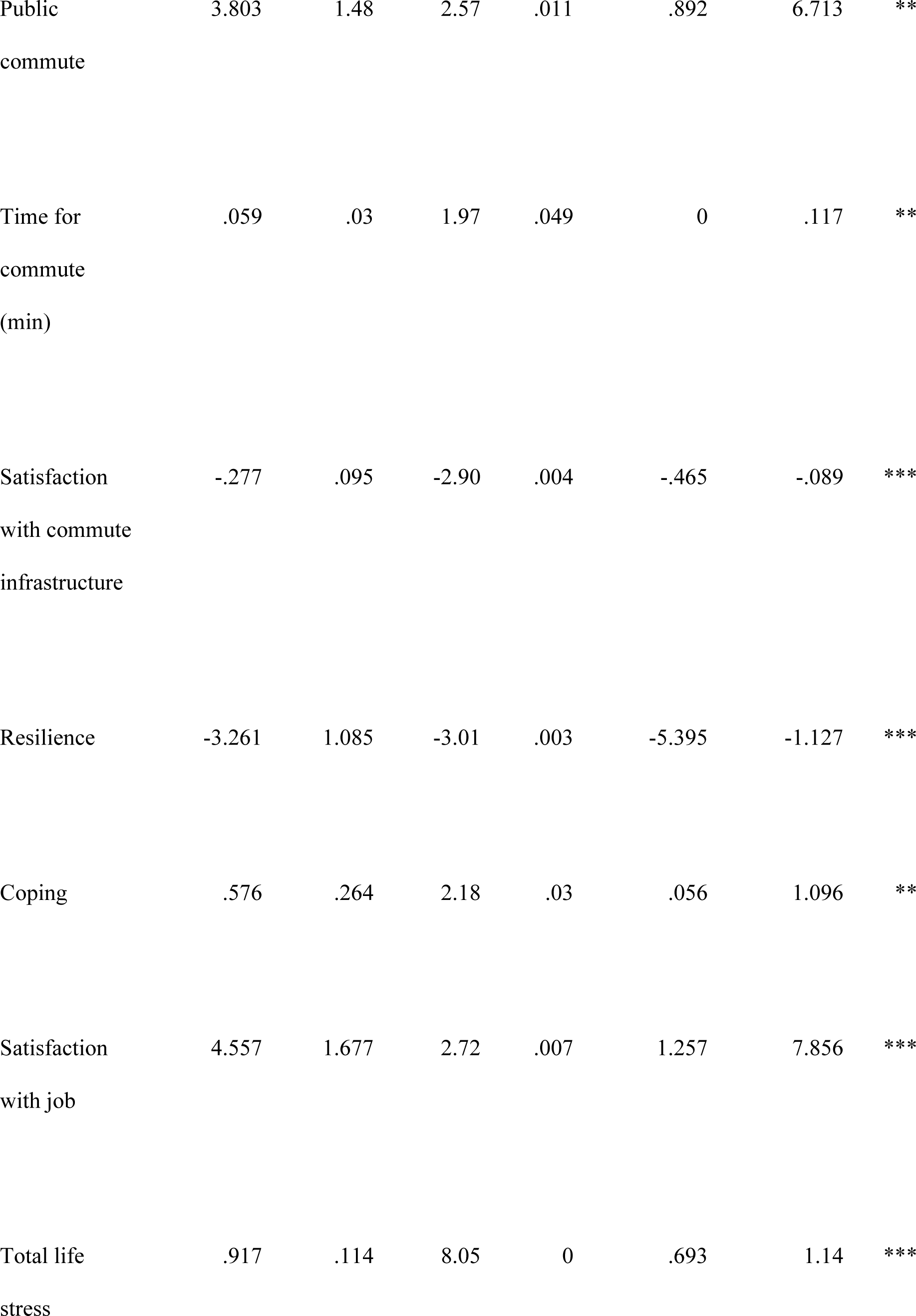
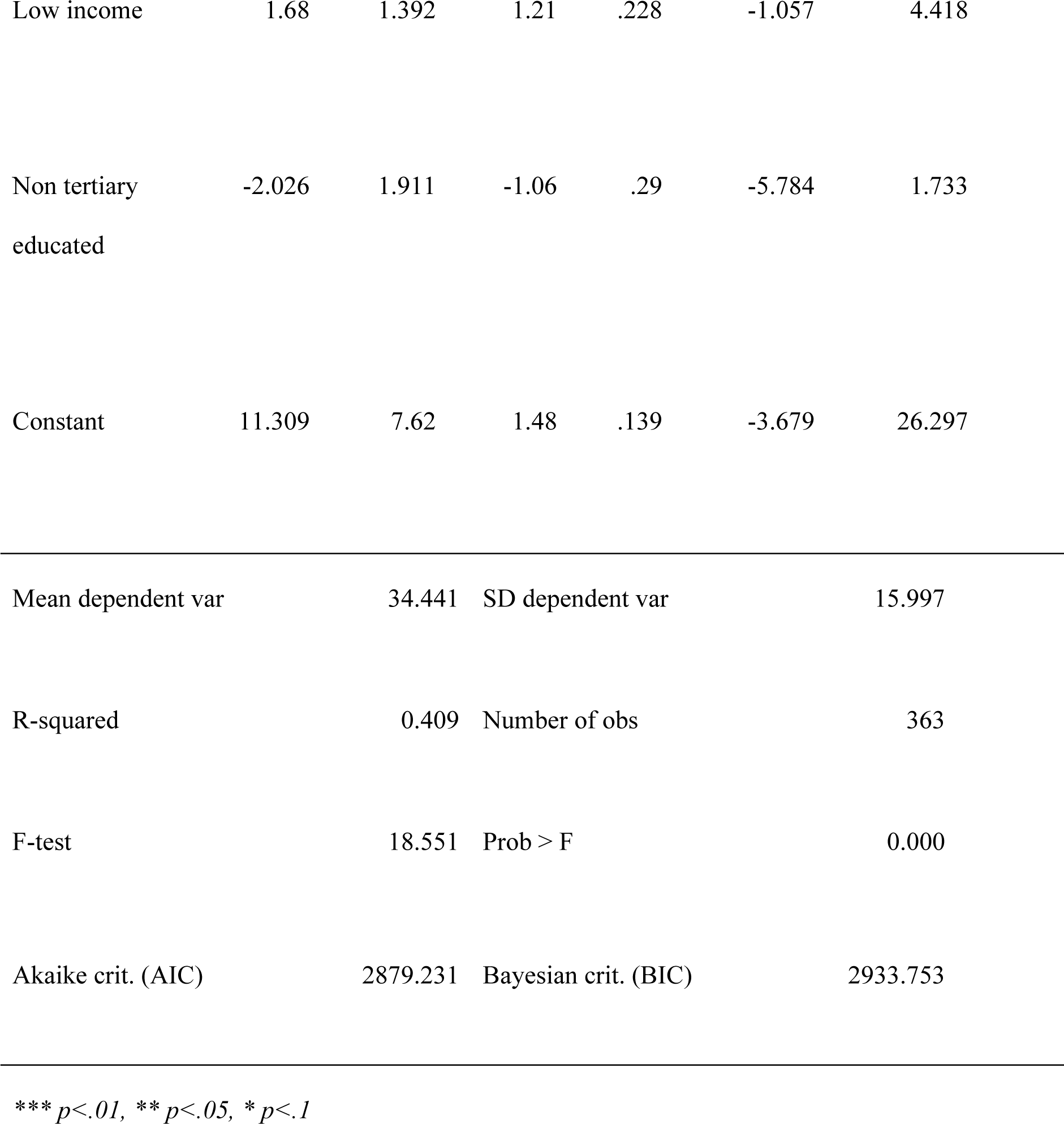
Effect of intrinsic and extrinsic factors on commuting stress score.

## Discussion

The main aim of this paper was to determine what factors were associated with experiencing commuter stress during the daily commute using symptomatology. After controlling for resiliency, coping, perception of job, total life stress, income levels and education all of which can contribute to how persons perceive hassles in their lives, the regression model demonstrated that females, persons who used public transportation, persons who have longer commute times and persons who are less satisfied with the commute infrastructure are more likely to report that they experience symptoms associated with stress while engaged in the daily commute compared to males, persons who use private transportation, persons who have shorter commute times and persons who are more satisfied with the commute infrastructure respectively. Once all controlled variables were accounted for, it appears that for this study population, age, where they live and whether or not they actively or passively commuted did not have any meaningful impact on whether they would report if they experienced stress associated symptoms while engaged in the daily commute. These findings are for the most part consistent with what is already published in the literature that used alternative methods of measuring commuting stress. In the present study, the non-association that was found between the active/non-active commuting and commuting stress was discordant with previous studies, however this could be explained due to sampling inefficiencies which resulted in the majority of the sample being users of non-active commuting modes.

The findings of this study are limited to persons who are employed with the University of Guyana or one of its affiliate organisations/institutions for at least 6 months and have to commute to work on a daily basis. Despite these limitations however, the findings of this study add to the existing knowledge in a few ways. Firstly, this appears to be the first time an attempt was made to examine all of the factors (both the intrinsic and extrinsic) that could potentially contribute to commuting stress in a single study. Previous research tended to only focus on a few of the factors and there was very little consistency across studies as to how commuting stress should be measured. Secondly, for the most part, commuting stress has been measured either by using electrodermal activity or a single Likert-point scale, however, this study used symptomatology. The only other study that used symptomology to assess commuting stress was conducted by Rezapour et al., (2021), however, the focus was narrow with respect to both the factors examined and the symptoms used when compared to the present study. Further, Rezaour treated each symptom as an individual outcome whereas in the present study, all of the symptoms were reduced to a single value that factored in any positive emotions that may occur on the daily commute to give a more holistic view of the stress experienced on the daily commute. The third way in which this paper contributes to the literature is the fact that symptomology could now potentially be used as an alternative method for measuring commuting stress as the findings of this study for the most part replicate that of what exists in the available literature. Furthermore, in doing so, this study strengthens the findings in the previously published studies as the findings are similar even when using a different method to measure the outcome variables. The fourth way in which this study contributes to the existing literature is that it expands the list of factors that can be considered when thinking about commuting stress. The two novel factors that emerged in this study are people’s perception of their jobs, their satisfaction with the commute infrastructure in place, and whether or not they commute via public or private means. The fifth and final way in which this paper contributes to the literature is that it adds to the geographical diversity in which this issue was examined. A cursory review of previous papers would reveal that the studies which have examined this issue have been skewed towards North America, Europe and Asia, where there has been heavy investment in public transportation, with little to no studies emanating from South and Central America, the Caribbean and Africa. Furthermore, the majority of studies that are available were conducted in economies that have completed the urban transition, that is more than 50% of their total population lives in urban areas. At the time of data collection, Guyana, located in South America is an economy in transition with less than 50% of its population living in urban agglomerations.

## Conclusions

We found that both the intrinsic and the extrinsic factors interact in complex ways to influence the overall stress experienced during commuting. Understanding these can help in designing interventions to make commuting a less stressful experience, whether it involves improving public transit systems, encouraging flexible work arrangements, or promoting stress management strategies for individuals. Interventions such as these can significantly reduce their exposure to stress which stands as an independent risk factor for CNCDs that continues to plague both the developed and developing world. As a policy response following three measures can be implemented. First, at a personal level, health care providers could be provided with some tools that they can use on their own to reduce stressful situations. For example, one could provide mindfulness - often used as a therapeutic technique to manage stress and promote mental wellbeing. It is a practice that involves focusing one’s awareness on the present moment, while calmly acknowledging and accepting one’s feelings, thoughts, and bodily sensations. [53–54]

The second, establishment of guidelines to ensure that the infrastructure that is being used by commuters is mentally friendly, refers to physical infrastructure designed and constructed with an understanding of the psychological and emotional impact it has on individuals who interact with it. It aims to create an environment that promotes mental well-being and reduces stress levels. City planners and transportation officials can create an infrastructure that supports mental well-being and reduces the stress associated with commuting. In the long run, mentally friendly infrastructure can contribute to happier, healthier, and more productive communities. This will require high-level interventions as there may be a need to make the public transportation system less chaotic, ensure that there is more greenery along major thoroughfares, and just simply ensure that the commuting infrastructure such as roads and footpaths are maintained.

The third and final way that is proposed to reduce the stress on the daily commute for organisations is to ensure that their employees are meaningfully engaged. An employee who is meaningfully engaged at work is deeply connected and committed to their role, their team, and their organisation. They find their work fulfilling and significant, feel that they are making a valuable contribution, and are motivated to do their best. To foster meaningful engagement, organisations need to provide supportive leadership, opportunities for growth and development, a positive work environment, and recognition for contributions and achievements. While everyone’s circumstances are different, and a range of factors can influence commute-related stress, a positive relationship with their work and their workplace can indeed help mitigate stress during their commute. For example, when employees find meaning in their work and feel valued in their roles, they may experience less anxiety about workplace challenges or pressures. This positive mindset can extend to their commute, making them less stressed about potential delays or complications on their way to work. Similarly, employees who are empowered to manage their workloads effectively may have more flexible schedules, allowing them to avoid peak commuting times or opt for less stressful modes of transport. In an organisation that nurtures meaningful engagement, the employee is likely to handle commuting stresses more effectively, viewing them as minor inconveniences which can contribute to an overall reduction in stress, both within and outside of work

## Data Availability

All relevant data are within the manuscript and its Supporting Information files.

